# Redefining Vasoplegia: A New Hemodynamic Model for Differentiating Cardiac vs. Vascular Dysfunction using the Resistance Flow Ratio

**DOI:** 10.64898/2026.01.28.26345090

**Authors:** Nikhil Patel, Menachem Weiner, Matthew A Levin

**Author notes:** **Corresponding Author Information:** – Matthew A Levin MD, Department of Anesthesiology, Perioperative and Pain Medicine, Icahn School of Medicine at Mount Sinai, New York NY 10029.

## Abstract

**Background:** Conventional definitions of vasoplegia and cardiogenic shock rely on fixed thresholds for mean arterial pressure (MAP), cardiac index (CI), and systemic vascular resistance (SVR), failing to account for the physiologic interdependence between cardiac output and vascular tone. We propose the *Resistance:Flow Ratio (RFR = SVRI/CI)* as a continuous, physiologically anchored measure to differentiate vascular versus cardiac dysfunction and to unify compensated, decompensated, and mixed shock states.

**Methods:** Single-center retrospective study of adult cardiac surgery cases, 2014-2024. Hemodynamic data for the first 72h post-operatively were analyzed. RFR thresholds were derived by modeling the relative efficiency of an increase in CI versus SVRI in improving perfusion pressure (PP: calculated as MAP minus central venous pressure) by 25%, identifying inflection points corresponding to vasoplegic, mixed, and cardiac-dominant physiology. Patients were categorized into six states by combining RFR-defined etiology with perfusion status (PP ≥50 vs <50 mmHg). Transition dynamics were analyzed using Markov chain modeling. Agreement with conventional definitions of vasoplegia and cardiogenic shock was assessed using sensitivity, specificity, predictive values, and accuracy relative to RFR-PP defined states.

**Results:** Out of 10,338 cases, 3,378 met inclusion criteria. RFR thresholds of <400, 400-900, and >900 corresponded to vasoplegic, mixed, and cardiogenic shock, respectively. Conventional (decompensated) vasoplegia occurred in 19.4% of patients, versus 22.8% by RFR-PP criteria, while 39.9% met criteria for compensated vasoplegia. Decompensations occurred within the same RFR category in 65% of cases, validating physiologic separation of etiology from compensation. Markov chain modeling revealed a postoperative drift from pump-failure to vasoplegic states during the postoperative course. RFR-PP definitions showed greater sensitivity than conventional criteria while maintaining high specificity.

**Conclusions:** The RFR-PP framework quantitatively separates cardiac from vascular dysfunction, captures compensated precursor states, and links directly to therapeutic logic. RFR-PP could provide a scalable platform for real-time, physiology-based hemodynamic assessment and shock management.

## Introduction

Differentiating hemodynamic compromise due to pathologic vasodilation (vasoplegia) versus primary pump failure (cardiogenic shock) is critically important in the management of cardiovascular decompensation following cardiac surgery. Traditional definitions of vasoplegia and cardiogenic shock, however, have a number of important limitations. They rely on fixed thresholds of mean arterial pressure (MAP), cardiac index (CI), and systemic vascular resistance (SVR), which fails to account for physiologic interdependence, where vascular resistance dynamically adjusts to changes in cardiac output.^1^ Recent systematic reviews also reveal that there is no consensus definition of vasoplegia, with heterogeneous MAP thresholds ranging from <50 to 80 mmHg and CI thresholds from >2.0 to 3.5 L/min/m².^2,3^ This leads to a wide variability in incidence and inconsistent outcome reporting.

Importantly, perfusion pressure (PP; MAP minus central venous pressure [CVP]), which has been shown to outperform MAP alone in predicting mortality and end-organ dysfunction, is not typically included in either the definition of vasoplegia or cardiogenic shock.^4^ These findings highlight the importance of integrating tissue perfusion into hemodynamic assessment. Another limitation is the use of SVR rather than indexed SVR (SVRI)^5^. Using SVR introduces Body Surface Area (BSA) as a confounder, potentially leading to systemic misclassification. Finally, mixed shock states (simultaneous cardiac and vascular dysfunction), which occur in a significant number of patients in cardiac critical care units, carry an elevated mortality risk yet are poorly captured by these binary classifications.

We propose the Resistance:Flow Ratio (RFR), defined as the ratio of SVRI to CI, as a more physiologically anchored descriptor of the balance between vascular tone and cardiac function. We use RFR in conjunction with perfusion pressure to create a comprehensive hemodynamic framework consisting of discrete hemodynamic states that map directly to therapeutic logic and capture transition dynamics after cardiac surgery. Combined, RFR plus PP overcome the limitations of the traditional definitions of vasoplegia.

## Methods

### Data Source

This retrospective study was approved by the Institutional Review Board of the Mount Sinai School of Medicine (IRB # STUDY-19-01166-CR004) with waiver of informed consent and all analyses adhered to ethical guidelines for the use of de-identified patient data. We extracted data for all adult patients undergoing cardiac surgery at a single high-volume urban academic medical center between January 2014 and December 2024. Sex– and race/ethnicity-stratified descriptive characteristics are reported, but subgroup analyses by sex or race/ethnicity were not assessed. Healthcare Cost and Utilization Project (HCUP)^6^ clinical classification software categories as well as current procedural terminology (CPT) codes were used to identify the type of procedure. Inclusion criteria comprised: patients aged ≥18 years who underwent (1) heart valve procedures (HCUP code 43), (2) coronary artery bypass grafting (HCUP 44), (3) aortic procedures requiring cardiopulmonary bypass (HCUP 52), (4) heart transplantation (CPT codes 33945, 33929, 33935), or (5) left ventricular assist device implantation (CPT codes 33979, 33981, 33980, 33983). Cases were required to have continuous hemodynamic monitoring data during the 72-hour postoperative period. We excluded patients with transcatheter procedures (e.g. TAVR, TEER), isolated mediastinotomy, percutaneous LVAD removal, and those missing measurements of MAP, CVP, or HR.

### Hemodynamic Data Collection & Temporal Aggregation

Hemodynamic parameters were extracted from the electronic medical record (EMR, Epic, Epic Systems, Verona WI) throughout the first 72 hours of the postoperative course, beginning from the documented case end time. In our institution, noninvasive measurements (e.g., HR, SBP/DBP, MAP, CVP) are captured every 15 minutes or hourly, whereas hemodynamic measurements derived from pulmonary artery catheters (CO/CI, SVR/SVRI) are typically recorded once per nursing shift. Individual patient hemodynamic profiles were constructed by binning the 72-hour post-operative period into 5-minute intervals, similar to the methodology described by Swan et al.^7^ Each interval could contain measurements for MAP, SBP, DBP, CVP, CO, CI, SVR, SVRI, and HR. When multiple MAP sources existed, the direct arterial line measurement was prioritized. Perfusion pressure was defined as MAP – CVP. CI was calculated as CO divided by body surface area (BSA), and SVRI as [(MAP – CVP) × 80] / CI. Measurements were assigned to the interval corresponding to their timestamp; no interpolation or carry forward was performed. Data cleaning and preprocessing included removal of nonnumeric entries, negative values, and physiologically implausible outliers using variable-specific thresholds (Supplemental table 1). The median measurement count per case and median time delay between measurements over the entire 72 hour post-operative period were calculated for each hemodynamic variable. Violin plots were constructed and visual inspection was used to determine the minimum measurement count threshold for inclusion. (Supplementary Figure 1) Hemodynamic variables were treated as a set; if the measurement count of any variable fell below the minimum, the case was excluded. This overall approach preserved data granularity while avoiding the artificial inflation of prevalence estimates that can arise from imputing missing values.

### Resistance-Flow Ratio (RFR) Threshold Determination

RFR was defined as the ratio of SVRI in dyn·s·cm⁻⁵·m² to CI in L/min/m² (equivalent to MAP-CVP*80/CI^2^). To establish physiologically relevant RFR thresholds, we analyzed the minimum independent percent increase from the mean baseline of either SVRI or CI required to achieve a clinically significant 10 mmHg increment in perfusion pressure, from a compromised perfusion pressure of 40 mmHg to a compensated PP of 50 mmHg. A threshold of 50 mmHg was chosen for two reasons. First, using a typical vasoplegia criteria of MAP < 60 mmHg and a typical normovolemic CVP of 8-12 (average 10) mmHg, a PP of less than 50 mmHg (60 – 10) would correlate to a hypoperfused, vasoplegic state. Second, existing literature suggests a wide range of minimum tissue PP from > 34 mmHg in septic shock and critical illness^8,9^, > 50 mmHg for hepato-splanchnic flow^10^, > 60 mmHg for prevention of acute kidney injury^11^, to 50-70 mmHg for cerebral perfusion in brain injury^10^. Together, these considerations justify 50 mmHg as a perfusion pressure threshold that extends the conventional MAP < 60 mmHg criterion while remaining aligned with physiologic tissue perfusion ranges reported across prior studies.

Two separate threshold points were identified where a 1.5-fold greater efficiency in increasing perfusion pressure by manipulating CI or SVRI was noted: a lower RFR threshold below which increasing SVRI was more efficient than increasing CI (indicating predominantly vascular dysfunction), and a higher RFR threshold above which increasing CI was more efficient than increasing SVRI (indicating primarily cardiac dysfunction). “Efficiency” was defined as requiring a smaller percent increase to achieve an equivalent gain in perfusion pressure. (full calculation in *Supplementary Figure 2*) The inflection points were rounded to the nearest 50 RFR units to improve bedside interpretability.

### Patient Categorization into a Unified Hemodynamic Framework

Perfusion pressure was used to determine whether a patient was hemodynamically compensated (PP ≥ 50 mmHg) or decompensated (PP < 50 mmHg). Patients were classified into six hemodynamic states by combining RFR-defined etiology (predominantly cardiac dysfunction, vascular dysfunction, or mixed) with perfusion status (compensated vs. decompensated), yielding a 3×2 matrix. The six states were: Compensated Heart Failure (CH), Decompensated Heart Failure (DH), Normal Function (N), Mixed Decompensation (DM), Compensated Vasoplegia (CV), Decompensated Vasoplegia (DV). Each timepoint with full hemodynamic data was classified independently, allowing for a single patient to occupy multiple states over time.

### Assessment of State Prevalence and Observed State Transitions using Markov Chain Modeling

To evaluate the clinical utility of the RFR-PP framework, we analyzed the prevalence and trajectory of hemodynamic states throughout the postoperative course and serial state classification was performed. Observed transitions between states were quantified, and transition matrices were constructed to describe the relative incidence of each type of clinical deterioration or recovery. Discrete-time Markov chain modeling was used to characterize the probabilistic dynamics of hemodynamic state transitions. Each state defined by the RFR-PP matrix was treated as a node in the chain, with transition probabilities derived from empirical frequencies across consecutive timepoints. A transition model was developed and stratified by initial postoperative state. The model was used to predict the evolution of hemodynamic states over time from initial postoperative distributions and to calculate steady-state distributions representing the long-term hemodynamic equilibrium. Predictions were then validated against observed temporal patterns in patient trajectories over the 72 hour postoperative course.

### Comparison to Conventional Definitions of Vasoplegia and Cardiogenic Shock

Finally, the newly proposed RFR-PP hemodynamic framework was compared to the gold standard conventional definitions of vasoplegia and cardiogenic shock. For vasoplegia, the traditional criteria of mean arterial pressure (MAP) <60 mmHg, cardiac index (CI) >2.2 L/min/m², and systemic vascular resistance (SVR) <800 dyn·s/cm⁵ were assessed against the novel RFR-PP based definition of decompensated vasoplegia (DV). For cardiogenic shock, the traditional criteria of MAP <60 and CI <2.2 L/min/m² were similarly assessed against the new definition of decompensated heart failure (DH). Conventional and RFR-PP-defined vasoplegia prevalences were calculated as the proportion of patients meeting all definition criteria in at least one timepoint. Using all available complete measurements sets from patients with PA catheters, confusion matrices were constructed to calculate sensitivity, specificity, positive predictive value (PPV), negative predictive value (NPV), and overall accuracy of traditional definitions relative to the RFR-PP definitions.

### Statistical Analyses and Computation

All data preprocessing, statistical analyses, and modeling were performed using Python (v3.9) and R (v4.2.2). Python libraries included pandas, NumPy, scikit-learn, matplotlib, and pyreadr; R packages included markovchain, ggplot2, pROC, and yardstick. Markov modeling and state-transition matrices were constructed in R with markovchain and visualized with ggplot2. We report means with standard deviations or medians with interquartile ranges, as appropriate. Two authors (NP, MAL) had full access to all the data and take responsibility for the integrity of the data and the accuracy of the analysis.

## Results

### Patient Demographics and Clinical Characteristics

The initial cohort comprised 10,338 cardiac surgery patients with basic hemodynamic monitoring during the postoperative course. After exclusion criteria were applied, 3,378 patients (32.7%) had sufficient PA catheter data for complete RFR analysis. The mean age was 63 (±12) years, with 66% male and 34% female. The cohort included a range of cardiac surgical procedures, including coronary artery bypass grafting (28.2%), heart valve (51.5%), heart transplant (7.3%), left ventricular assist device (10.4%), and aortic procedure with bypass (2.5%). Full demographic and clinical characteristics are presented in Table 1.

**Table 1:**
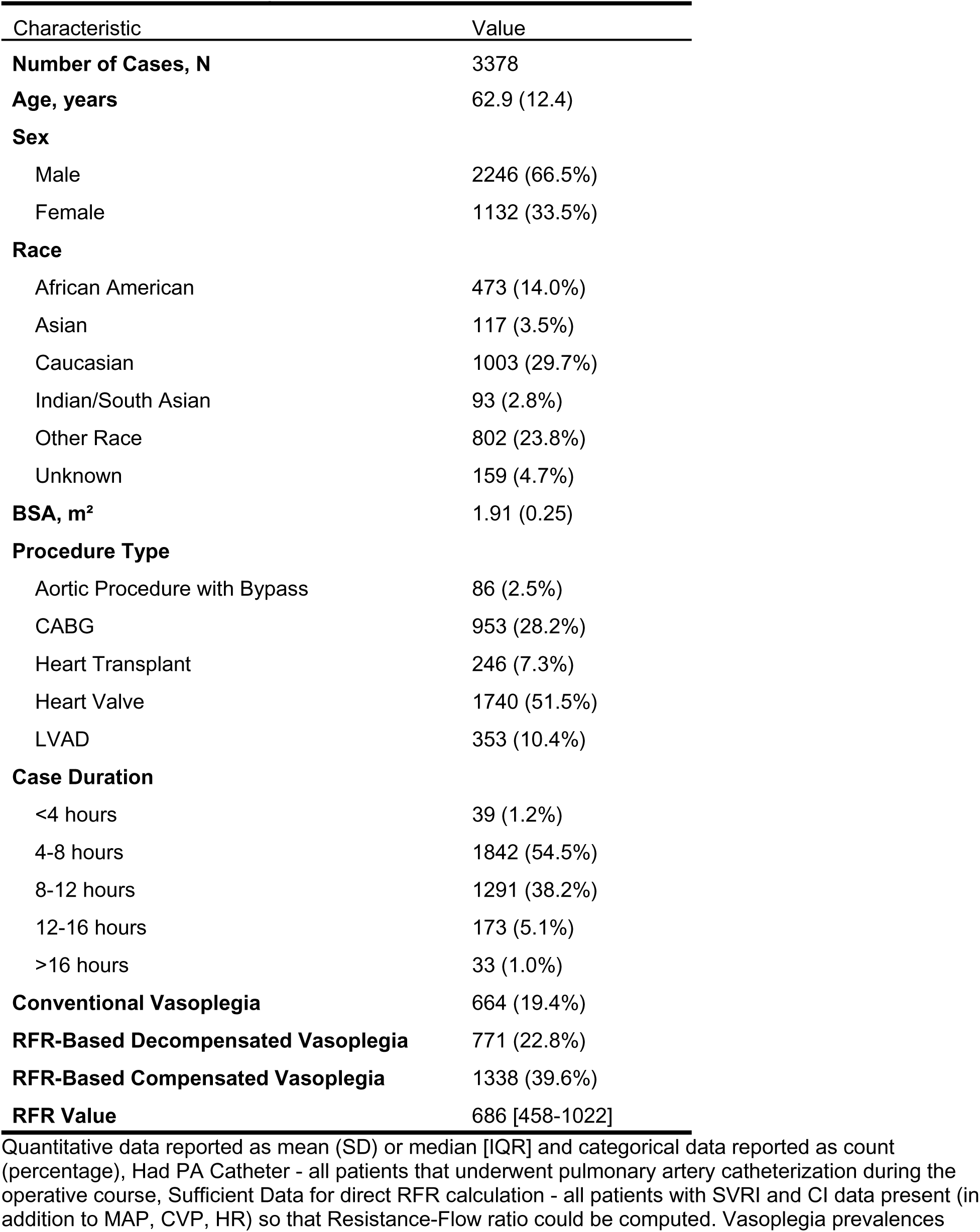
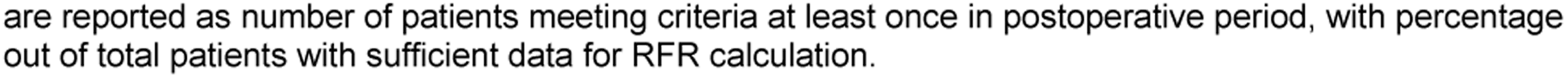
Cohort Demographic and Clinical Characteristics.

**Table 2:** Hemodynamic States in RFR-based Model.

| RFR | Interpretation | Compensated | Decompensated | Treatment |
| --- | --- | --- | --- | --- |
| > 900 | **Cardiac output optimization** is $\geq 1.5\times$ more efficient than vasoconstriction to increase perfusion | Compensated Heart Failure | Decompensated Heart Failure | Inotropes |
| 400–900 | Cardiac output optimization and vasoconstriction have **near equal efficiency** in increasing perfusion | Normal Function | Decompensated Mixed Etiology | Combo |
| < 400 | **Vasoconstriction** is $\geq 1.5\times$ more efficient than cardiac output optimization to increase perfusion | Compensated Vasoplegia | Decompensated Vasoplegia | Pressors |

MAP, SBP, and DBP were co-recorded a median of 111 times per case (median interval between successive measurements 39 minutes). CVP was recorded 77 times (median interval 56 minutes), and HR 98 times (median interval 44 minutes) (Supplemental Figure 1). Given these measurement frequencies, < 25 measurements for core variables (MAP, SBP, DBP, HR, or CVP) was chosen as the threshold for exclusion. After exclusion, hemodynamic parameters demonstrated near normal distributions, indicating a representative sampling of the overall cardiac surgery population (Supplemental Figure 3).

The mean number of complete simultaneous measurements (HR, MAP, CVP, CI, and SVRI) per case was 6.5, corresponding to one full set approximately every 12 hours across the 72-hour postoperative window (Supplemental Figure 4). Mean (SD) values were: HR 93.2 (16.6) bpm, MAP 76.5 (9.4) mmHg, CVP 12.0 (5.1) mmHg, CI 2.8 (0.8) L/min/m², and SVRI 1968.7 (696.9) dyn·s·cm⁻⁵·m² (Supplemental Figure 3). The median RFR value was 685.5 with an interquartile range of 458 to 1022. (Supplemental Figure 3).

### Threshold Determination for Resistance-Flow Ratio (RFR)

We identified inflection points at which cardiac output optimization versus vasoconstriction provided ≥1.5x efficiency over the other in raising PP 10 mmHg. These points corresponded to an RFR > 900, favoring increased CI (minimum 16% CI increase vs. 24% SVRI increase required, indicating predominant cardiac dysfunction), and RFR < 400, favoring increased SVRI (minimum 16% SVRI increase vs. 24% CI increase required, indicating predominant vasoplegia). Intermediate values between 400 and 900 were defined as representing mixed states with similar efficiency between cardiac output optimization and vasoconstriction. *Figure 1* shows a modeling plot generated to visualize this efficiency relationship across a physiologic range of SVRI and CI.

**Figure 1:**
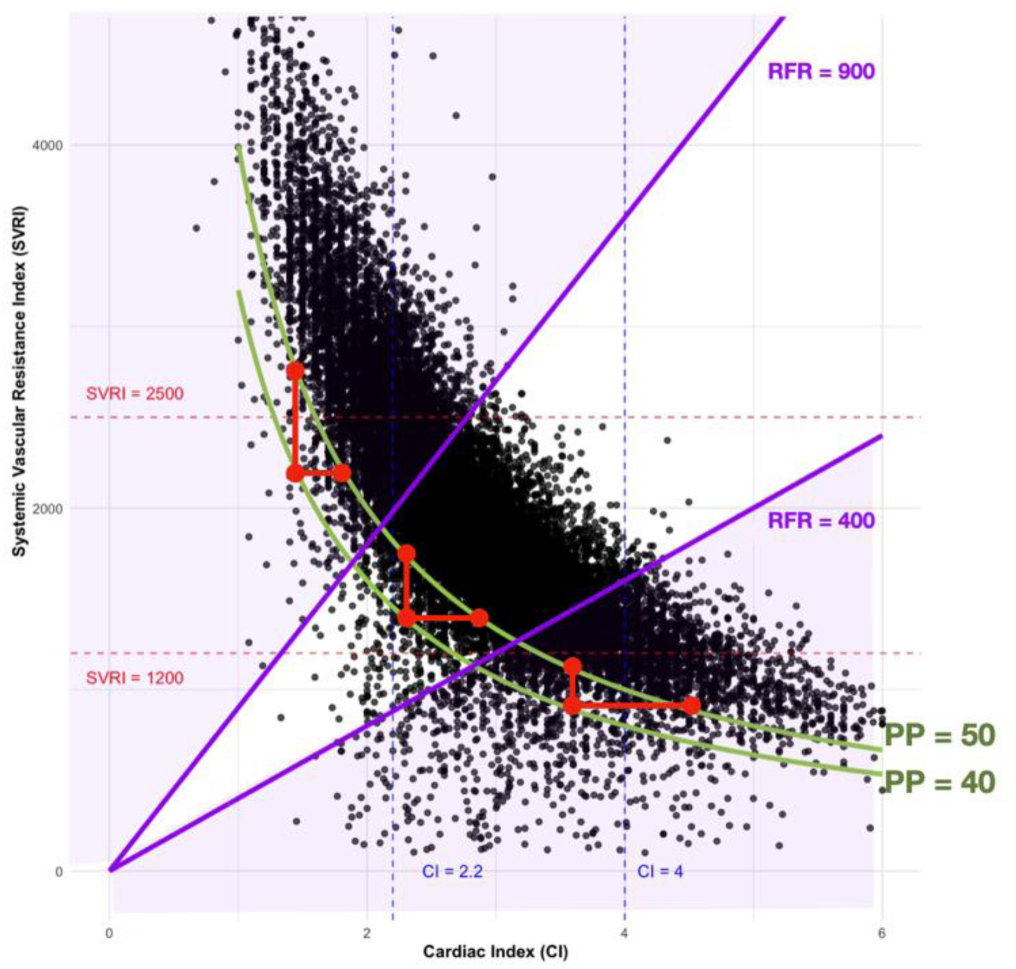
Efficiency Graph for RFR Threshold Determination. Scatter plot showing minimum increases in SVRI or CI required to raise perfusion pressure by 10 mmHg (red lines). Green curves = PP levels (40, 50). Purple lines = RFR thresholds. Black points are observed measurement pairs of CI and SVRI in the clinical cohort.

### Prevalence of Hemodynamic States

Patients were categorized into the six distinct hemodynamic states in *Table 1* using the identified RFR thresholds: 1) compensated heart failure (RFR>900, PP>50), 2) decompensated heart failure (RFR>900, PP<50), 3) normal function (*N*: RFR 400-900, PP>50), 4) mixed decompensation (RFR 400-900, PP<50), 5) compensated vasoplegia (RFR<400, PP>50), and 6) decompensated vasoplegia (RFR<400, PP<50). Correspondence of hemodynamic states with observed measurements of CI and SVRI are visualized in *Figure 2*.

**Figure 2:**
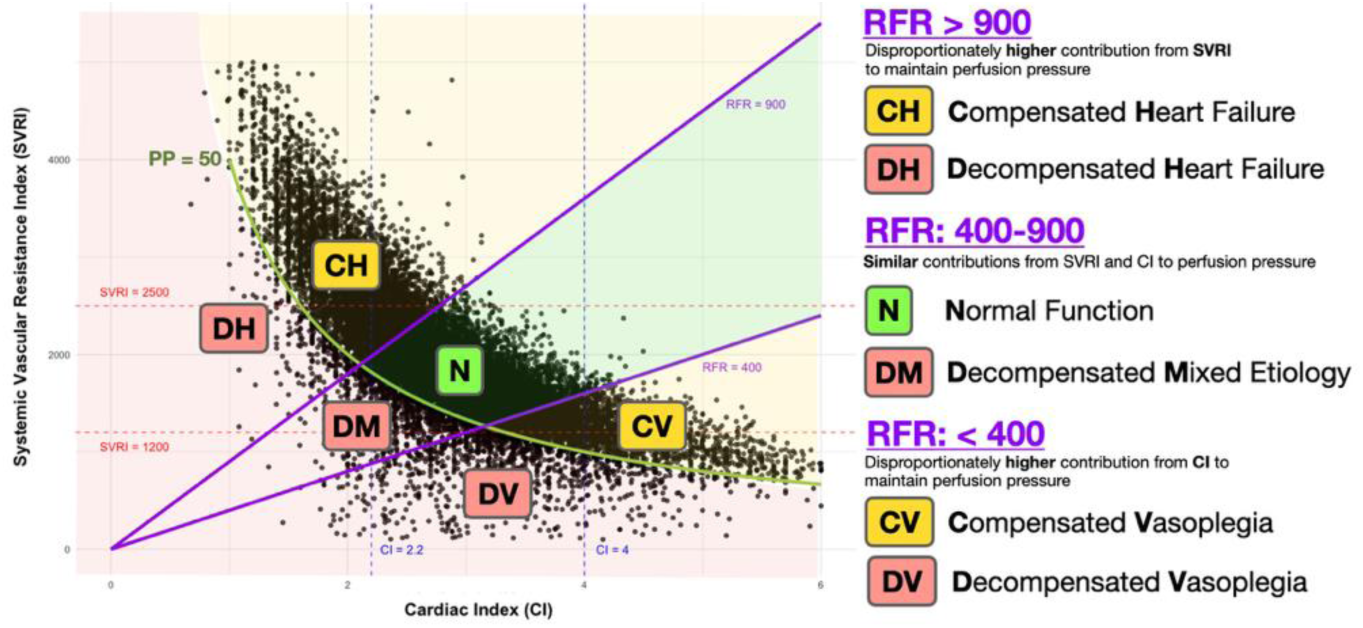
SVRI vs. CI Scatter Plot Visualization of RFR-based States. SVRI vs. CI scatter plot with regions defining six states. Green curve = critical PP (50 mmHg), determining compensation status. Purple lines mark RFR (SVRI/CI) categories (>900, 400-900, <400).

Most patients (87.7%) exhibited normal function at least once during the post-operative period, yet 39.6% of patients met the criteria for compensated vasoplegia at least once, 22.8% qualified as having decompensated vasoplegia at least once, and nearly a fifth (19.4%) of patients appeared to be in a mixed decompensated state. (Supplemental Figure 5)

### Observed State Transitions

Analysis of serial timepoints revealed the characteristic transitions between hemodynamic states shown in *Figure 3*. The most frequent transitions were between compensated and normal states. Among transitions from a compensated/normal to a decompensated state, most (65%) occurred within the same RFR category (pump failure, normal/mixed, or vasoplegia). The most common decompensation was to a mixed decompensated state (862/1,877 transitions, 46%). 63% of decompensations were from a compensated state compared to 37% from normal functioning. Notably, decompensation into lower (vasoplegic) RFR states occurred more than twice as frequently as decompensation into higher (cardiogenic) RFR states, with 373 vs. 151 transitions, respectively. Overall flux during the post-operative course was toward lower more vasoplegic RFR states (3,748 transitions) versus higher more cardiogenic RFR states (2,973 transitions).

**Figure 3:**
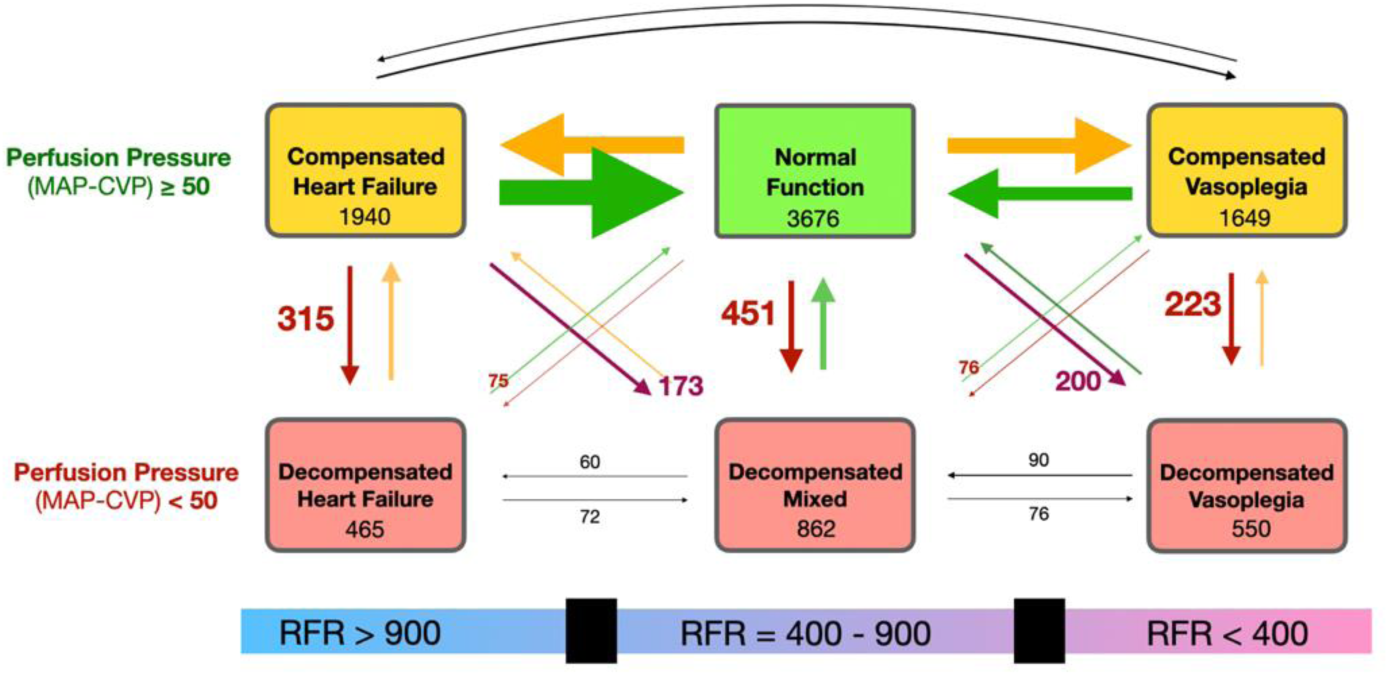
Hemodynamic State Transition Diagram. Hemodynamic State Transition Diagram. Box numbers indicate total transitions into each state (incidence). Arrows represent transitions, scaled by transition frequency. Numbers adjacent to arrows show transitions into decompensated states.

### Markov Chain Model Analysis

Markov chain modeling accurately predicted the evolution of hemodynamic states toward steady-state equilibrium as depicted in *Figure 4*. The model predicted notable population shifts from initial postoperative distributions, including decreasing prevalence of compensated heart failure (–34.8%) and decompensated heart failure (–49.0%) as well as increasing prevalences of normal (+21.2%) compensated vasoplegia (+48.0%), mixed decompensation (+15.2%) and decompensated vasoplegia (+16.2%). These model predictions closely align with observed patient trajectories over 72 hours postoperatively, with particularly notable increases in vasoplegic states and decreases in heart failure states after the first 24 hours post-surgery.

**Figure 4:**
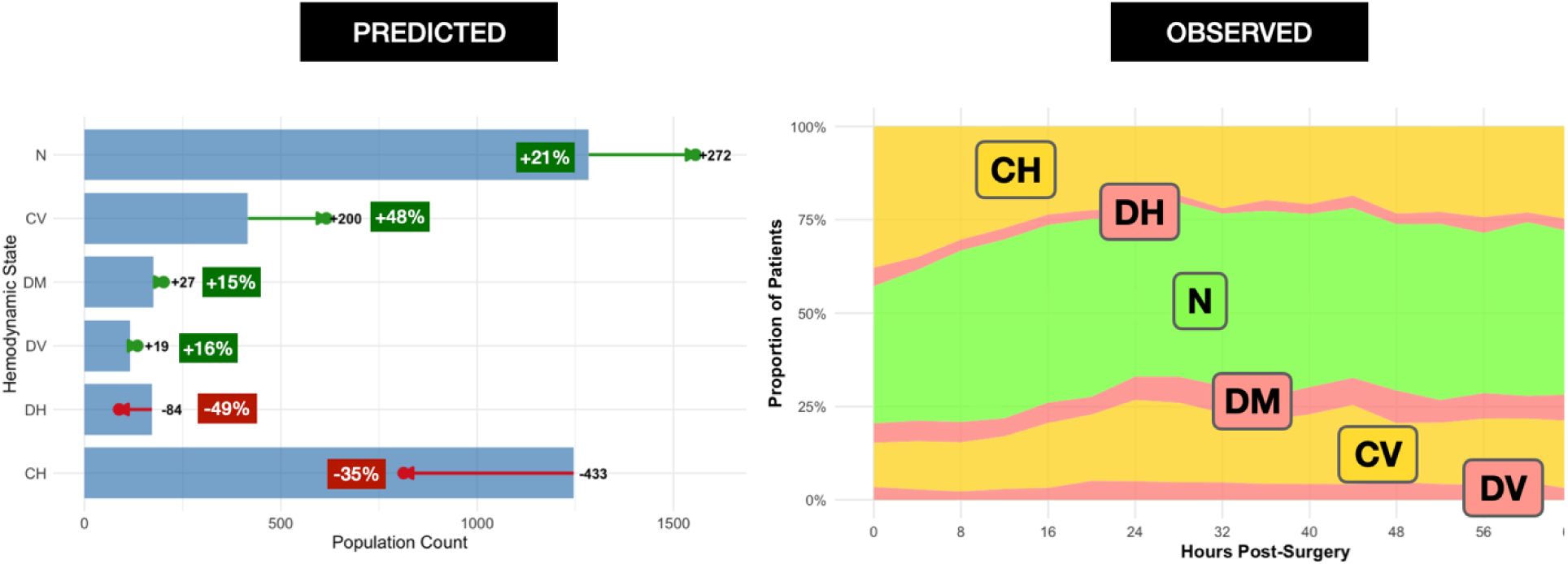
Markov Chain Prediction vs Observed Postoperative State Progression. Left: Predicted postoperative progression to steady state by Markov chain modeling. Blue bars represent initial populations immediately following surgery. Arrows indicate the predicted magnitude and direction of changes. Predicted percentage change is also displayed. Right: Observed postoperative state proportions over 60 hours. States include compensated heart failure (CH), decompensated heart failure (DH), normal function (N), mixed (DM), compensated vasoplegia (CV), and decompensated vasoplegia (DV).

### Concordance with Conventional Definitions of Vasoplegia and Cardiogenic Shock

Conventionally defined vasoplegia (MAP < 60 mmHg, CI > 2.2 L/min/m², SVR < 800 dyn·s/cm⁵) occurred at least once during the postoperative course in 19.4% of patients, whereas RFR-PP defined decompensated vasoplegia occurred in 22.8% and RFR-PP defined compensated vasoplegia in 39.6% of patients. RFR-PP defined compensated and decompensated heart failure occurred in 64% and 7% of patients, respectively. Reasonably strong concordance was observed between traditional definitions of cardiogenic shock and vasoplegia and the newly established RFR-PP classifications for both decompensated heart failure and decompensated vasoplegia, with high specificity (99.4% and 98.8%, respectively) and accuracy (98.5% and 96.6%). However, sensitivities were considerably lower at 27.1% for decompensated heart failure and 57.7% for decompensated vasoplegia. The reduced sensitivity was primarily attributable to a substantial number of false negatives, driven by conventional definitions’ reliance on MAP rather than perfusion pressure. This resulted in conventional definitions excluding patients who exhibit hypoperfusion despite having a preserved MAP. NPVs were high for both definitions, at 99.1% for decompensated heart failure and 97.7% for decompensated vasoplegia, while PPVs were lower at 34.8% for decompensated heart failure and 72.5% for decompensated vasoplegia, reflecting the presence of false positives. False positives were predominantly due to conventional definitions categorizing hemodynamic states as cardiogenic shock or vasoplegia when, according to the RFR-PP model, these cases represented mixed decompensation rather than simple isolated DH or DV. Confusion matrices, statistics, and scatterplots comparing definitions are detailed in *Figure 5*.

**Figure 5:**
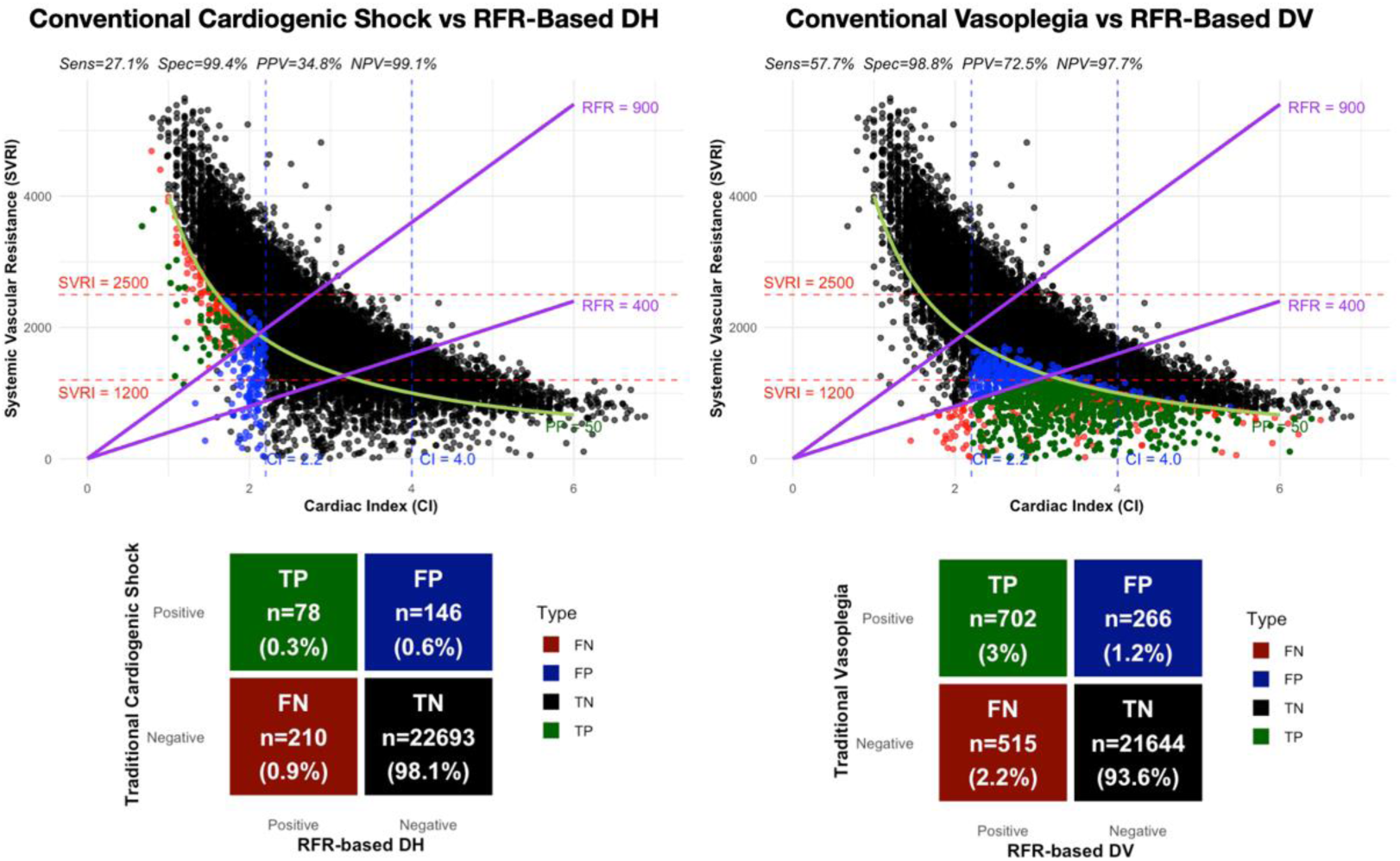
Comparison of RFR-based Definition to Conventional Definitions of Vasoplegia and Cardiogenic Shock. CI vs SVRI scatterplots (top) with colored points corresponding to confusion matrices (bottom), where TP = True Positive (green), FP = False Positive (blue), FN = False Negative (red), TN = True Negative (black). Left side compares conventional cardiogenic shock definition to RFR-based decompensated heart failure (DH, treating as ground truth). Right side compares conventional vasoplegia definition to RFR-based decompensated vasoplegia (DV, treating as ground truth)

## Discussion

In this large, contemporary cohort study of cardiac surgical patients, we found that the RFR, a new framework for categorizing post-cardiac surgery shock, effectively discriminates between cardiogenic and vasoplegic shock, while providing new insight into mixed shock states. The incidence of decompensated vasoplegia by RFR criteria closely matched that of the conventional definition of vasoplegia, yet almost 40% of patients met RFR criteria for being in a compensated vasoplegic state, with increased cardiac output but poor vascular tone. The RFR integrates systemic vascular resistance index and cardiac index into a single physiologic construct that faithfully stratifies the full spectrum of postoperative hemodynamics. Low RFR denotes vasoplegia; high RFR indicates pump failure; intermediate values capture mixed states. Because it is continuous, RFR supports trending with response assessment, and paired with perfusion pressure, it separates compensated from decompensated presentations.

Across prior work, SVRI and CI have almost always been handled as separate axes used to plot phenotypes, set quadrant cut-points, or qualitatively label “high/low” combinations, rather than being collapsed into a single scalar that discriminates vascular from cardiac dysfunction. A prior study of 31 patients described phase-dependent changes in the SVRI & CI relationship during the inflammatory response, underscoring the relevance of evaluating these parameters jointly in hemodynamic assessment.^12^ In congenital Fontan physiology, Kawasaki et al. profiled patients on a CI-SVRI plane similar to our framework and defined four hemodynamic categories, but did not combine the terms into a ratio or produce a continuous index of etiology.^13^ Their follow-up similarly categorized patients based on quadrants and demonstrated prognostic separation without a unifying ratio.^14^ Others have modeled the CO-SVR manifold itself using a nonlinear regression (CO ≈ 38.5 × SVR^-0.799) that reproduces the Ohm’s law perfusion pressure curve. They defined “afterload-related cardiac performance (ACP)” as the vertical deviation from a tolerance band, which helps quantify cardiac impairment at a given afterload, but does not distinguish vascular from cardiac drivers.^15^ Acute heart failure phenotyping has used cardiac power index (CPI = MAP x CI) and drawn empirical zones based on identified cases of cardiogenic shock, CHF, and septic shock on CPI-SVRI plots similar to our CI-SVRI plot; however, the authors acknowledge these boundaries are arbitrarily drawn.^16^ CPI also conflates reduced MAP from vasodilation with reduced CI from pump failure, limiting utility when vascular dysfunction coexists. Unlike CPI plots (which conflate MAP with CI) or ACP deviations (which quantify pump function at a given afterload while ignoring the vascular contribution), RFR directly partitions vascular versus cardiac drivers and identifies the most efficient bedside lever (tone vs inotropy).

Beyond cardiology, cirrhosis work has compared CO-SVR curves by ascites severity and demonstrated a left-shift (lower perfusion pressure) in refractory ascites, but did not index for BSA or analyze ratio directionality, leaving the vascular vs cardiac contribution unresolved.^17^ In emergency department sepsis, unsupervised clustering on noninvasive SVRI and CI produced three phenotypes with markedly different 30 day mortality; however no differences in clinical features were identified and no scalar discrimination of etiology was provided.^18^ Notably, the two low-CI clusters from this study are most consistent with cardiac-dominant and mixed sepsis-cardiogenic phenotypes rather than pure distributive shock, aligning with their higher 30-day mortality; in our RFR-PP framework, these map to high-RFR states. Finally, outpatient hypertension work used the inverse ratio (CI/SVRI) to describe chronic hemodynamic phenotypes; associations with demographics were weak, the context was stable hypertension rather than shock physiology, and there was no linkage to perfusion pressure or acute therapeutic guidance.^19^

Examining the behavior of this new definition in practice, five findings with immediate clinical implications emerged. First, an RFR of 400-900 appears to delineate the “physiologic set point” at rest; excursions above or below this window mark compensated precursor states that precede overt shock in > 60% of deteriorations, underscoring a pre-shock interval for preventative therapy. Second, the observation that most decompensations occur within the same RFR category (e.g. compensated vasoplegia to decompensated vasoplegia) supports the identified thresholds and the separation of etiology from compensation. Third, because each state is anchored in the relative efficiency of manipulating cardiac output versus vascular tone, this framework maps directly onto treatment logic: inotrope-predominant for high-RFR states, vasoconstrictor-predominant for low-RFR states, and dual or device-based strategies for mixed states. Compared with conventional binary definitions that ignore venous congestion and treat cardiogenic and distributive shock as mutually exclusive, the RFR-perfusion-pressure matrix captures covert hypoperfusion, quantifies mixed phenotypes, and improves sensitivity without sacrificing specificity, thereby unifying disparate guideline concepts into a single actionable schema.

Fourth, longitudinal mapping showed a drift from pump-failure phenotypes (high RFR) toward vasoplegic phenotypes (low RFR) within the first 24 hours after surgery, followed by progressive stabilization, providing insight into a biphasic postoperative course. To our knowledge, this is the first quantitative, time-resolved description of a biphasic postoperative course in adults; prior pediatric studies noted an early CI nadir/SVRI peak with subsequent recovery but neither framed an etiologic drift from pump-failure to vasoplegia nor modeled state transitions.^20,21^ Our RFR-PP based Markov analysis reveals delayed vasoplegia emergence as cardiac function and MAP improve (↑CI → ↑MAP → baroreflex-mediated ↓SVR ± vasoplegia mechanisms), implying that vasopressor needs may rise even as output normalizes, necessitating proactive titration.

Finally, our incidence estimates reinforce external validity. Using direct PA-catheter measurement, the RFR-PP definition identified vasoplegia in 23% compared with 20% by conventional criteria among 3,378 patients, indicating modestly greater sensitivity; These figures sit well within prior literature showing a mean incidence 19.9% (95% CI 16.1-24.4%) across 60 studies in a 2025 systematic review^2^ and 5-25% in an earlier reviews.^22^ Notably, this cohort constitutes, to our knowledge, the second-largest cohort with a directly measured vasoplegia incidence.

### Established Clinical Shock Presentations Correspond to RFR-Defined States

Cardiogenic shock has traditionally been described using a 2x2 phenotype matrix classifying patients as ‘warm’ or ‘cold’ based on peripheral perfusion, and ‘wet’ or ‘dry’ based on volume status.^23^ The RFR-PP states map directly onto these phenotypes. Specifically, decompensated heart failure encompasses both the ‘cold and wet’ and ‘cold and dry’ phenotypes, thereby capturing hypervolemic as well as euvolemic cardiogenic shock presentations. The integration of perfusion pressure rather than a fixed mean arterial pressure (MAP < 60 mmHg) threshold ensures identification of hypervolemic ‘cold and wet’ patients who may otherwise be missed by conventional rule-based definitions (e.g., patients with normal MAP but significantly elevated CVP causing hypoperfusion). Additionally, mixed decompensation aligns with the ‘warm and wet’ phenotype, and decompensated vasoplegia corresponds to the ‘warm and dry’ phenotype, covering the spectrum of shock presentations.

### RFR Establishes Specific Criteria for Recognition of Compensated Precursor States

The introduction of the RFR offers a novel methodology for identifying compensated precursor states, such as compensated vasoplegia (CV) and compensated heart failure (CH), which are not quantitatively defined in current ACC/AHA guidelines. Existing guidelines characterize high-output heart failure and distributive pre-shock states primarily through clinical features and broad hemodynamic trends including warm extremities, strong pulses, and preserved mentation rather than numerical measures.

For example, high-output heart failure is described in the setting of elevated cardiac output (typically >8 L/min) and low systemic vascular resistance, but without strict cutoff values for diagnosis^1,24^. Similarly, pre-shock distributive states are acknowledged by preserved or elevated cardiac output and reduced SVR, yet no numeric thresholds for SVR or CI are provided^1,25^. In contrast, the RFR framework establishes precise, empirically derived thresholds that reflect hemodynamic imbalance, potentially enabling identification of patients in compensated vasoplegic and compensated pump failure states, which are unrecognized by guideline criteria focused on overt hypotension or shock. As compensated states most often deteriorate into decompensated states, this early recognition is critically important.

### Limitations

Several limitations merit consideration. The dataset is retrospective and single-center, derived largely from cardiac-surgical patients with PA catheters, and therefore subject to selection bias, institutional practice patterns, and potential measurement error in derived variables. The selection of a 50 mmHg perfusion pressure threshold and a 10 mmHg delta PP, while physiologically reasoned and supported by prior literature, remains partly arbitrary. Similarly, the efficiency-based RFR cut-points are empiric, and both elements may require refinement with broader outcome-based validation.

### Conclusion

The RFR is a novel, simple metric that effectively differentiates between vasoplegic and cardiogenic shock, while also identifying mixed shock states. Prospective, multicenter studies including patients with non-surgical shock will be essential to refine RFR thresholds, confirm generalizability, and test whether RFR-guided algorithms can improve time to appropriate therapy, organ-function recovery, and survival.

## Data Availability

A deidentified copy of the data analyzed will be made available upon reasonable written request.

## Acknowledgments

We used an assistive writing tool to help with language editing. All authors reviewed the content and take full responsibility for the integrity and originality of the manuscript.

## Materials and Data Availability

De-identified data underlying this study and associated analysis code are available from the corresponding author upon reasonable request and completion of a data use agreement, as approved by the institutional review board.

## Non-standard Abbreviations and Acronyms

ACP: Afterload-Related Cardiac Performance
BSA: Body Surface Area
CABG: Coronary Artery Bypass Grafting
CH: Compensated Heart Failure
CI: Cardiac Index
CO: Cardiac Output
CPB: Cardiopulmonary Bypass
CPI: Cardiac Power Index
CPT: Current Procedural Terminology
CV: Compensated Vasoplegia
CVP: Central Venous Pressure
DH: Decompensated Heart Failure
DM: Mixed Decompensation
DV: Decompensated Vasoplegia
EMR: Electronic Medical Record
HCUP: Healthcare Cost and Utilization Project
LVAD: Left Ventricular Assist Device
MAP: Mean Arterial Pressure
ML: Machine Learning
N: Normal Function
NPV: Negative Predictive Value
PA: Pulmonary Artery
PP: Perfusion Pressure (MAP – CVP)
PPV: Positive Predictive Value
RFR: Resistance–Flow Ratio
SBP: Systolic Blood Pressure
SVR: Systemic Vascular Resistance
SVRI: Systemic Vascular Resistance Index

## Supplemental Material

**Supplementary Table 1:** Hemodynamic Outlier Criteria.

| Parameter | Acceptable Range (Outliers outside of this range removed) |
| --- | --- |
| SVRI | 200-6000 |
| CI | 0.5-8 |
| MAP | 30-150 |
| CVP | 0-30 |
| HR | 20-250 |

**Supplementary Figure 1:**
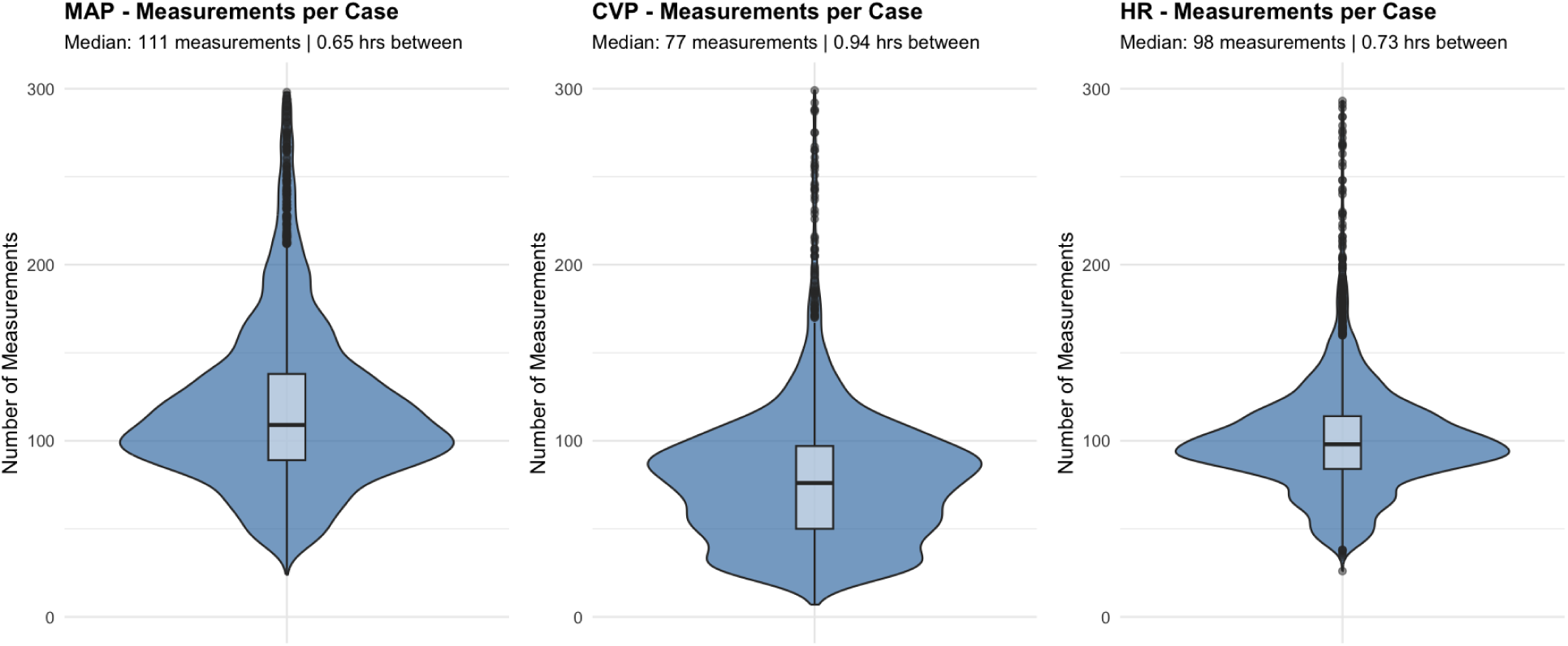
Distributions of Measurement Counts. *SBP and DBP have the same measurement frequencies as MAP as they are co-measured

**Supplementary Figure 2:** Calculation of Relative Efficiency.

The relative efficiency of cardiac index (CI) compared to systemic vascular resistance index (SVRI) in increasing perfusion pressure from 40 to 50 mmHg was calculated using the following formula:

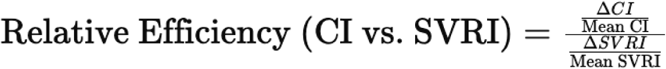

Where:

- ΔCI = Independent increase in CI required to raise perfusion pressure from 40 to 50 mmHg
- Mean CI = 3.1 L/min/m²
- ΔSVRI = Independent increase in SVRI required to raise perfusion pressure from 40 to 50 mmHg
- Mean SVRI = 1850 dyn·s/cm⁵/m²

**Supplementary Figure 3:**
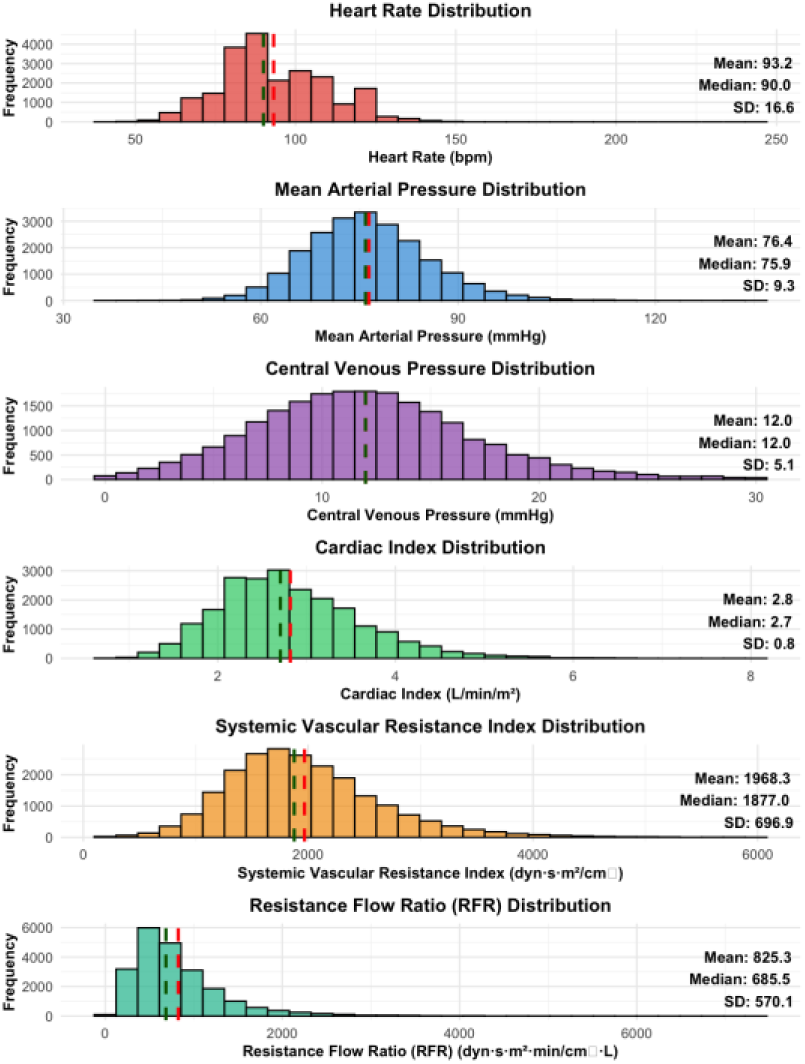
Distributions of Measured Hemodynamic Parameters in PA Catheter Cohort. Histograms of measurements of MAP, CVP, HR, CI, SVRI, and RFR in patients with PA Catheters (n = 3378)

**Supplementary Figure 4:**
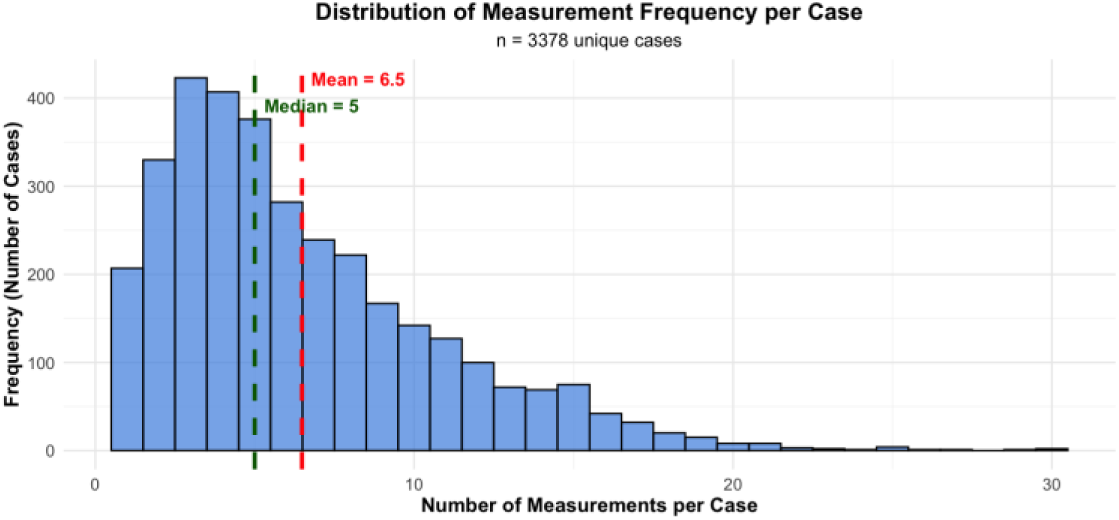
Complete Measurement Frequency. Number of Complete Measurements Sets (HR, CI, SVRI, CVP, MAP) per case (n = 3378)

**Supplementary Figure 5:**
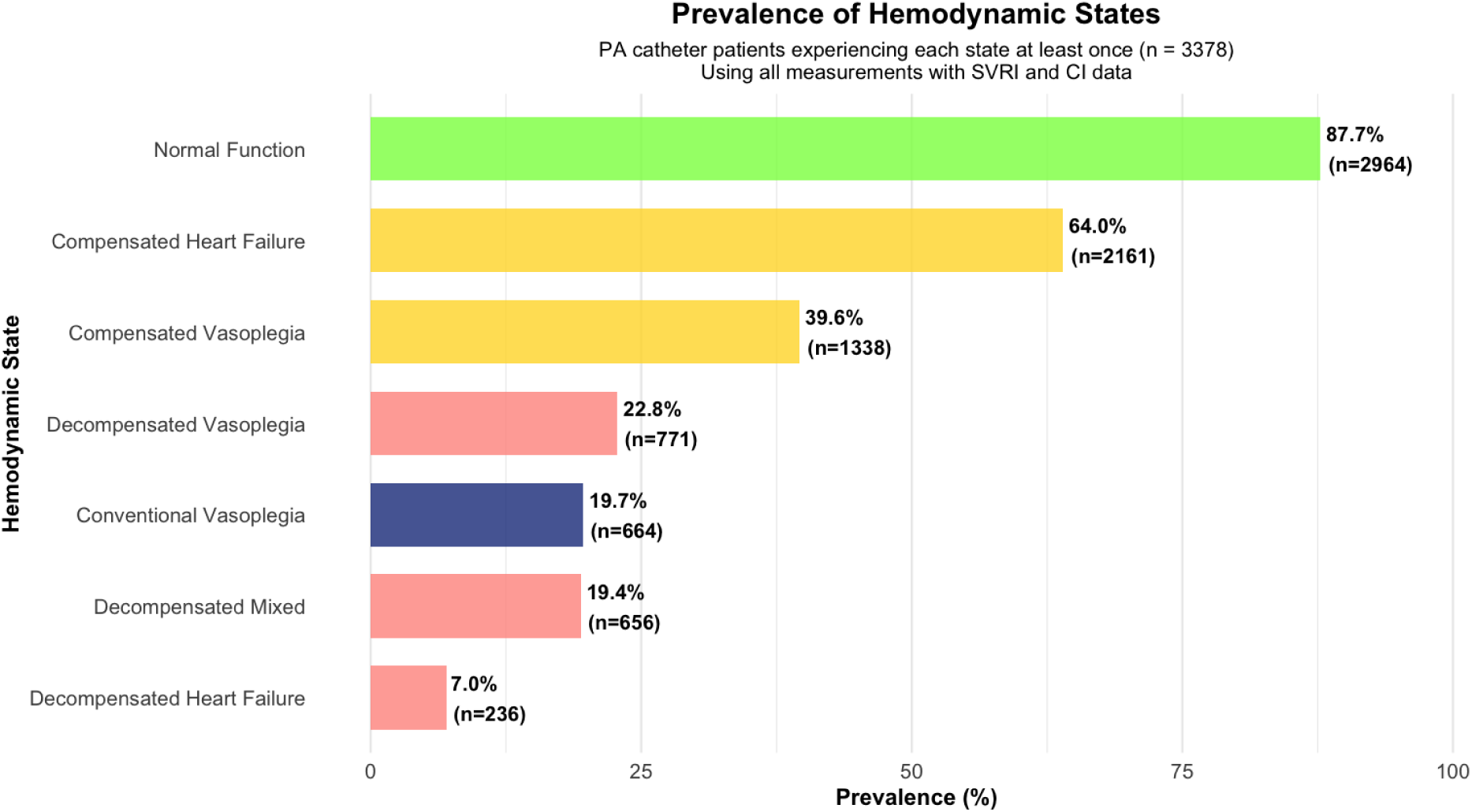
Prevalence of All Hemodynamic States. Patients experiencing each state at least once during postoperative course in patients with PA Catheters (n = 3378)

